# CuGen: A GPU-accelerated framework for large-scale genomics

**DOI:** 10.64898/2026.07.15.26358178

**Authors:** Tuomo Kiiskinen, Joshua Richland, William Wang, Sophia Lu, Balasubramanian Narasimhan, Trevor Hastie, Robert Tibshirani, Manuel A. Rivas

## Abstract

Biobank-scale genomic analyses remain computationally expensive, CPU-bound workflows, particularly when adjusting for confounding. Here, we present CuGen, a GPU-accelerated framework for large-scale genomics. CuGen uses UltraLasso, a novel hierarchical application of univariate-guided sparse regression (uniLasso), to select a compact, phenotype-informed active set of fewer than 30,000 variants. This achieves robust leave-one-chromosome-out (LOCO) confounding control, enabling both downstream GWAS and in-sample fine-mapping. Additionally, we introduce the `.cugen` file format, a genotype representation designed for memory-optimized, high-throughput streaming and random access on GPU hardware. Building on this substrate, we provide a general GPU-accelerated genomics toolkit handling polygenic prediction, data manipulation, quality control, analysis, and visualization. We demonstrate CuGen’s efficacy in the UK Biobank with up to 408,624 individuals, where the full GWAS pipeline and fine-mapping against 6.8 million imputed variants completes in approximately 10 minutes on a single high-throughput GPU with 80 GB of memory. The pipeline scales efficiently to massive phenome-wide analyses with sublinear resource consumption.

## 1 Introduction

Genome-wide association studies (GWAS) have grown from small candidate-locus analyses into an enterprise of enormous computational scale [1, 2]. Modern biobanks routinely measure hundreds of thousands of individuals at millions of variants [3– 5]. Because the additive model evaluates variants independently, a GWAS requires solving millions of linear regressions relating phenotype to genotype while adjusting for covariates:

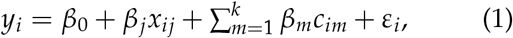

for each of *p* variants measured across *n* individuals.

The independence of these tests creates analytical challenges driven by covariance. Linkage disequilibrium between neighboring variants requires fine-mapping to resolve true causal signals from broader haplotype associations. Furthermore, individuals are correlated through shared ancestry and cryptic relatedness, which inflates test statistics if left unadjusted. Linear mixed models (LMMs) mitigate this confounding using a random effect built from an *N* × *N* genetic relationship matrix (GRM) [6–9]. Alternatively, whole-genome regression frameworks like REGENIE achieve similar control using leave-one-chromosome-out (LOCO) residuals from a fitted whole-genome predictor [10].

Because GWAS involves millions of independent calculations, it is an ideal candidate for massive parallelization. However, the sheer volume of tests far exceeds the thread count of modern CPUs, rendering biobank-scale analyses sequential workloads that run for days. Graphics processing units (GPUs) excel at applying single instructions across multiple data elements concurrently, yet their adoption in large-scale genomics has been hindered by two major bottlenecks. First, modern confounding control is memory-bound; LMMs and dense whole-genome ridge regressions construct dense objects, such as an *N* × *N* GRM or predictors spanning up to a million variants, that easily exceed the 40 to 80 GB capacity of modern GPUs. Second, genomic data movement is heavily I/O-bound. When the full genotype matrix cannot fit into device memory, the constant transfer of data between the host and the GPU dominates the runtime, nullifying the compute advantage.

Here we present CuGen, a GPU-native framework that restructures biobank-scale genomics to overcome these memory and I/O barriers. Cu-Gen’s statistical core is UltraLasso, a hierarchical application of univariate-guided sparse regression (uniLasso) [11]. Previously, we demonstrated that highly sparse polygenic risk scores built with uniLasso, comprising fewer than 40,000 active variants, achieve predictive accuracy matching state-of-the-art dense methods that utilize over ten times as many predictors [11, 12]. Here, we harness this extreme sparsity for confounding control in GWAS. By applying uniLasso’s penalized statistical learning algorithm in a hierarchical, stepwise fashion, UltraLasso isolates a highly compact, phenotype-informed active set. This achieves robust LOCO confounding control entirely within 80 GB of GPU memory. Finally, the pipeline completes with maximally parallelized association testing directly followed by UltraSuSiE—a GPU-native, in-sample application of Sum of Single Effects fine-mapping. Rather than treating fine-mapping as a disconnected post-GWAS analysis reliant on summary statistics and external reference panels, CuGen ties causal inference directly to the primary association scan. By performing Bayesian fine-mapping against the exact same dependent variable—the LOCO residual—used in the original association test, UltraSuSiE eliminates both reference panel LD mismatch and the analytical discrepancies of post-hoc inference [13–15]. To feed this architecture without I/O throttling, we introduce the `.cugen` file format, a CUDA-optimized genotype representation [16] designed for sustained streaming and constant-time random access.

We validate this framework using both polygenic simulations and real-world analyses in the UK Biobank, leveraging up to 408,624 individuals and 6.8 million imputed variants. Compared directly against REGENIE, CuGen delivers comparable or superior confounding control while condensing the full analytical pipeline—from sparse model construction and association testing to genome-wide fine-mapping—into approximately ten minutes on a single high-throughput GPU (NVIDIA H100) with 80 GB of memory.

## 2 Results

### 2.1 The UltraLasso GWAS framework

Biobank-scale genome-wide association studies in CuGen are driven by UltraLasso, a GPU-native statistical learning algorithm designed for confounding-controlled association testing (Fig. 1). Like established methods such as REGENIE, UltraLasso utilizes leave-one-chromosome-out (LOCO) predictions to account for confounding from population structure and relatedness. However, rather than relying on a dense whole-genome ridge regression across all array variants, UltraLasso introduces a hierarchical application of uniLasso [11, 12]. This sparse, univariate-guided regression algorithm produces sign-consistent models requiring substantially fewer active predictors to adequately capture the genetic background.

**Figure 1.**
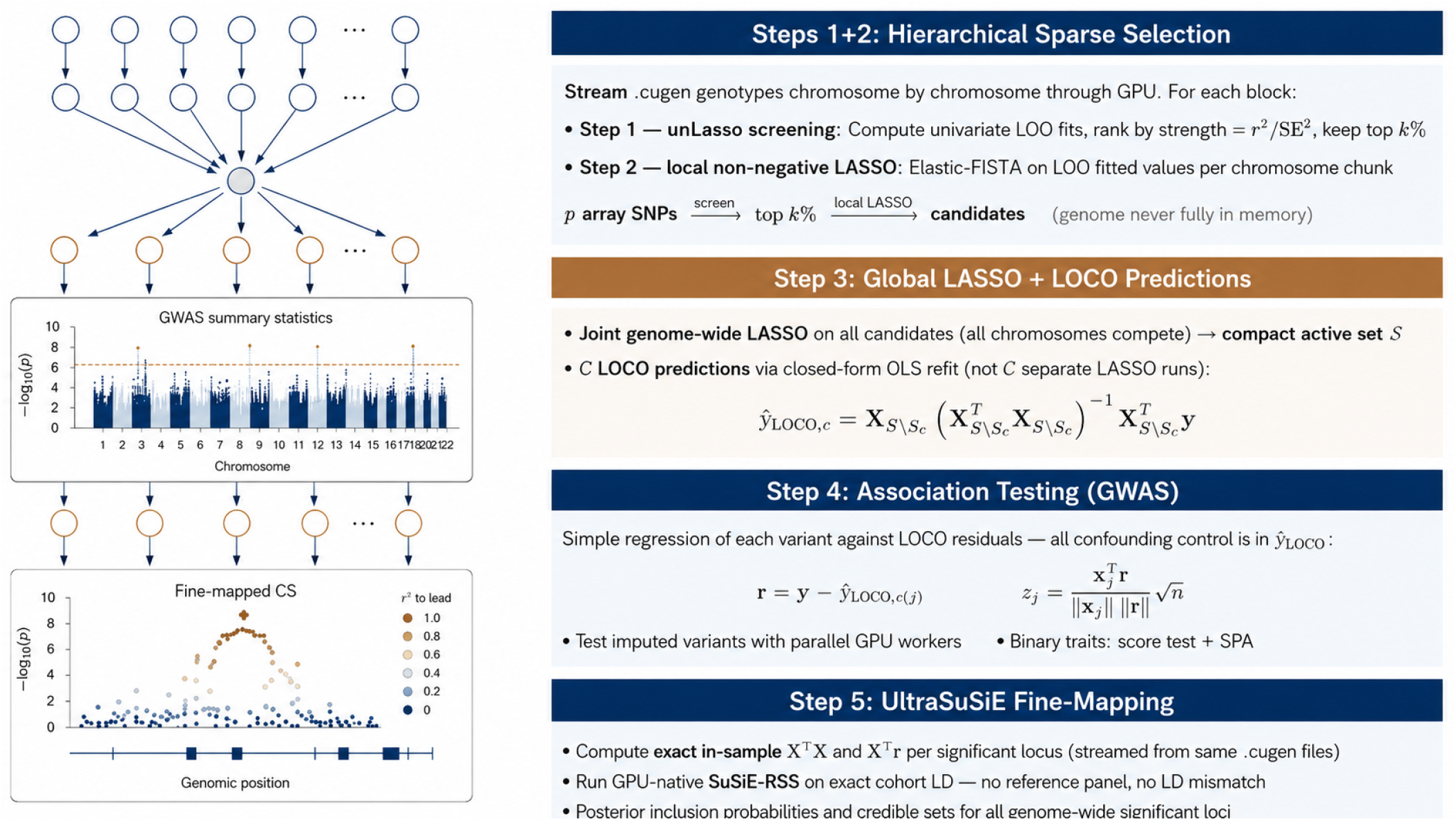
Overview of the CuGen UltraLasso GWAS framework. Array genotypes are streamed from .`cugen` files into GPU memory and passed through univariate screening and hierarchical sparse regression (uniLasso) to construct a compact active set, from which leave-one-chromosome-out (LOCO) predictors are built. Association testing regresses each imputed variant against the LOCO residual, and genome-wide significant loci are passed to UltraSuSiE for Bayesian fine-mapping with exact in-sample linkage disequilibrium. Benchmarks were run in the UK Biobank white British cohort with relatives excluded (*n* = 336,442) and included (*n* = 408,624).

To achieve this sparsity at scale, array genotypes are streamed chromosome by chromosome. Within each genomic chunk, the algorithm applies two stages of feature selection: a univariate leave-one-out fit ranks variants by a strength-based statistic, followed by a local non-negative LASSO where surviving variants compete. After processing the full genome, the candidates from each chunk are aggregated into a joint genome-wide LASSO. This final selection step yields a highly compact active set, typically comprising 10,000 to 30,000 variants. For each chromosome, a LOCO predictor is constructed via a closed-form ordinary least squares (OLS) refit on the active set that incorporates a minimal ridge penalty and explicitly excludes variants located on the target chromosome to prevent proximal leakage.

Downstream association testing is then reduced to a simple regression of each imputed variant against these LOCO residuals, a highly parallelizable task distributed across multiple GPU workers.

For binary phenotypes, the pipeline leverages a linear probability model coupled with a correction for case-control imbalance, maintaining computational speed without sacrificing statistical calibration.

Every step of this construction was cheap in compute and fit within the 80 GB device. On standing height in the full cohort, screening a chunk cost roughly 0.1–0.3 s of GPU compute, and each local non-negative LASSO—a 1,000-iteration FISTA over a few thousand candidates—took about 1–4 s. The single expensive step, the joint genome-wide LASSO, ran a 1,000-iteration FISTA over 38,455 candidate variants in 52 s after a 25 s precomputation of **X**^⊤^ **X** and **X**^⊤^ **y**, with the design matrix occupying 62.9 GB and leaving roughly 21 GB of headroom on the device. The 22 leave-one-chromosome-out OLS refits together took 12 s (about 0.5 s each).

Two elements of this construction proved necessary for numerical stability. The joint genome-wide LASSO cannot be bypassed: feeding the pooled local-LASSO candidates directly into the LOCO refits—a far larger and more collinear predictor set—produced unstable fits, whereas pruning to the joint active set first did not. And the minimal ridge in the per-chromosome OLS refits (*ε* = 10^−2^; Methods) is what keeps the occasionally near-singular Gram submatrices well conditioned.

### 2.2 The .cugen format enables GPU-native genotype I/O

Once the sparse model eliminates the GPU memory barrier, genotype data movement becomes the primary computational bottleneck. The .`cugen` (Compute Unified Genomes) file format is designed to resolve this constraint (Fig. 2). Standard genomic formats, such as PLINK .bed and .pgen, are optimized for CPU-oriented random access. They utilize sample-major or mixed layouts, lack pre-computed per-variant statistics, and rely on memory-mapped input/output routines that severely thrash the page cache during the long sequential scans required by genome-wide association studies. To accommodate GPU architecture, the .`cugen` format stores genotypes in a variant-major layout with 2-bit packing, ensuring each variant occupies a contiguous byte range tailored for coalesced device reads. Additionally, pre-computed per-variant statistics live directly in the file header. This enables on-device standardization within a single fused kernel and allows fixed-size records to support constant-time random access without requiring an external index.

**Figure 2.**
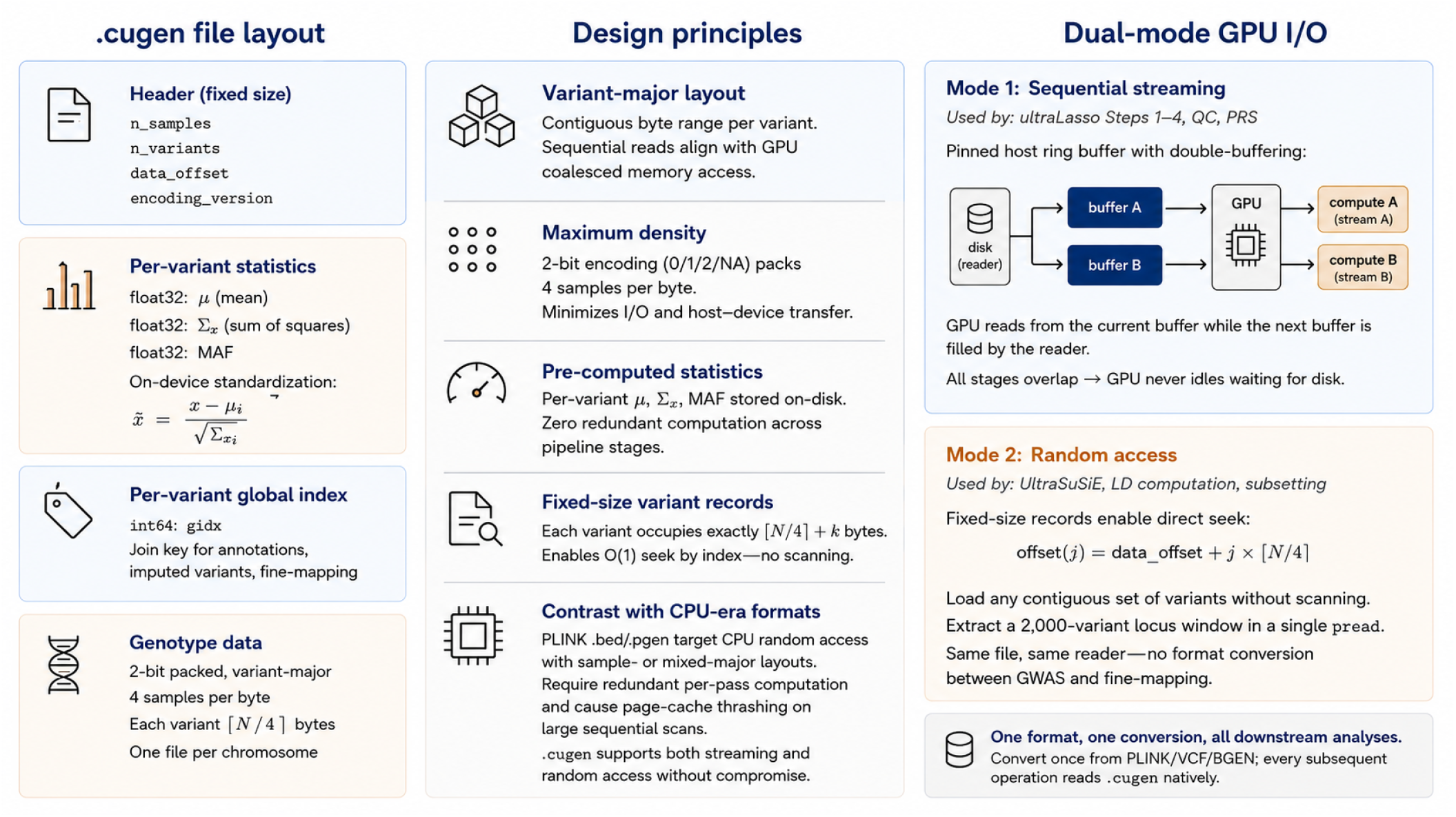
The .cugen (Compute Unified Genomes) file format. **a**, File layout: a fixed-size header storing pre-computed per-variant statistics (*µ*_*x*_, *S*_*xx*_, minor-allele frequency) and a global variant index, followed by 2-bit-packed, variant-major genotype data. **b**, Design principles: variant-major layout for coalesced GPU reads, fixed-size records enabling *O*(1) random access, and header statistics that eliminate redundant per-pass computation. **c**, Dual-mode GPU I/O: sequential streaming with pinned double-buffering for whole-genome scans, and direct random access for locus-level operations such as fine-mapping.

This data representation supports two distinct operational modes from a single file. In streaming mode, genotypes are continuously read into a pinned host ring buffer, asynchronously transferred to the device, and unpacked natively on the GPU. This ensures that disk reads, transfers, and computations overlap seamlessly, preventing the GPU from idling. Alternatively, the fixed record layout enables a random-access mode where any contiguous variant set can be extracted directly, supporting downstream tasks like in-sample fine-mapping and linkage disequilibrium computation. While this framework does not yet implement a full direct-to-GPU storage pipeline, the multi-worker streaming approach accelerates genotype reading by more than 20-fold compared to standard memory-mapped formats (Supplementary Table S1). Ultimately, this format conversion is a one-time, per-biobank cost: once the .`cugen` files exist, all further subsetting, quality control and analysis run as fast GPU-native operations with no additional data-movement penalty. Converting the roughly one million array variants of the full UK Biobank cohort (*N* = 408,624) from PLINK .pgen to `.cugen` took 21 minutes, split across 22 parallel CPU jobs.

### 2.3 Sparse LOCO residualization controls confounding and captures more phenotypic signal

Sparse LOCO residualization effectively controls confounding from population structure and relatedness across biobank-scale datasets. Using standing height as a primary quantitative benchmark in the UK Biobank, UltraLasso controlled confounding as effectively as dense whole-genome regression while capturing substantially more of the post-covariate phenotypic signal. We analyzed both an unrelated white British cohort (*N* = 336,442) and the full cohort including relatives (*N* = 408,624). The model was highly parsimonious: in the full cohort an active set of 22,301 variants reached an in-sample *R*^2^ of 0.76 for standing height (covariates and SNPs jointly), and this sparse fit proved stable—the 22 per-chromosome LOCO refits closely bracketed the full-model *R*^2^ (Supplementary Table S2). After covariate adjustment, the sparse LOCO predictor captured approximately half of the remaining phenotypic variance in each cohort (Supplementary Table S3).

Calibration under the null remained well controlled even when close relatives were included in the analysis. Across 200 null simulations (100 per cohort), the genomic inflation factor *λ*_GC_—the ratio of the median observed association *χ*^2^ statistic to its expectation under the null, equal to 1.0 in the absence of inflation—was at target for common variants (*λ*_GC_ = 1.000±0.005 in the full cohort at MAF ≥ 0.01; Supplementary Figs. S1–S2 and Supplementary Table S4). In polygenic simulations comprising 1,000 causal variants, UltraLasso recovered over 74% of causal loci at a false discovery rate near 10% in both cohorts (Supplementary Table S5). The preservation of confounding control after retaining approximately 72,000 relatives demonstrates that sparse LOCO residualization successfully absorbs cryptic relatedness without requiring a dense mixed model.

The framework also explicitly prevents proximal leakage. Repeating the entire variant selection independently for each held-out chromosome produced z-scores nearly identical to the production run (*r* = 0.997, median |Δ*z*| = 0.03; Supplementary Fig. S3). Furthermore, variants neighboring the active SNPs were no more discordant with REGENIE than frequency- and effect-size-matched non-neighbors (median|Δ*z*| 0.379 versus 0.380; Supplementary Fig. S4). This confirms that a shared active set combined with a closed-form OLS refit safely isolates the test chromosome, avoiding the signal entanglement seen in naive coefficient-subtraction methods.

When compared directly with REGENIE on matched cohorts, UltraLasso produced highly concordant genome-wide statistics, nearly identical at the aggregate level (Fig. 3), and comparable LD Score regression (LDSC) confounding ratios— the share of test-statistic inflation attributable to confounding rather than polygenic signal, for which lower is better—of 8.6–9.6% versus 9.4–9.8% (Supplementary Table S6). UltraLasso’s per-variant statistic, a standardized regression score, agrees with a per-variant Wald test to within numerical precision on the real height scan (Supplementary Table S7). However, the sparse LOCO predictor captured roughly 1.5 times the post-covariate variance using approximately 33 times fewer variants (Supplementary Fig. S5a). By removing more nuisance variance from the residual, this enhanced capture directly translates to statistical power. On identical simulated phenotypes, UltraLasso detected significantly more causal loci than REGENIE at a matched false discovery rate across all allelefrequency strata (paired *t*-test *P* = 3.2 × 10^−7^; Supplementary Fig. S5b, Supplementary Table S8).

**Figure 3.**
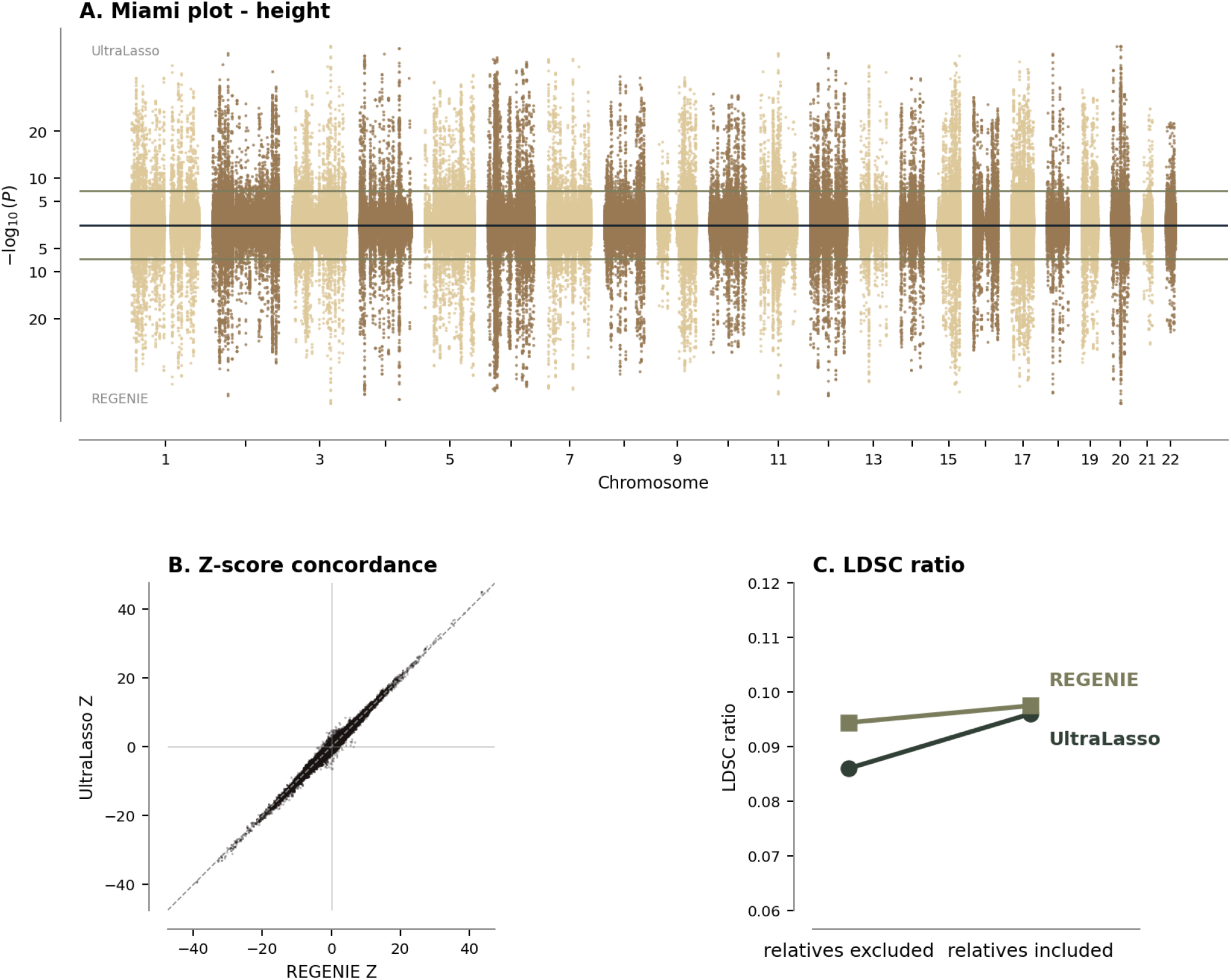
Concordance between UltraLasso and REGENIE for standing height. **a**, Miami plot contrasting Ultra-Lasso (top) and REGENIE (bottom) genome-wide association statistics in the full white British cohort (*n* = 408,624). **b**, Per-variant *z*-score concordance between the two methods. **c**, LDSC confounding ratios for both methods in the unrelated (*n* = 336,442) and full (*n* = 408,624) cohorts.

### 2.4 The framework extends to binary traits

The same sparse LOCO framework extends to binary traits through a single logistic recalibration. We applied it to asthma (54,901 cases), coronary heart disease (CHD; 17,111 cases), and type 2 diabetes (T2D; 25,144 cases) in the full cohort (*N* = 408,624), testing imputed variants at MAC ≥ 20. All computationally expensive fitting remains on the linear scale: the 0/1 phenotype is residualized by a linear probability model, the initial feature selection and genome-wide LASSO run as for a quantitative trait, and the chromosome-specific LOCO offsets are recalibrated on the logistic scale before a score test with selective saddlepoint approximation (Methods). Across all three traits, the recalibrated pipeline produced well-calibrated summary statistics with the expected polygenic tails (Fig. 4): asthma yielded 13,960 genome-wide significant variants (*λ*_GC_ = 1.174); CHD yielded 677 significant variants (*λ*_GC_ = 1.006); and T2D yielded 3,655 significant variants (*λ*_GC_ = 1.155).

**Figure 4.**
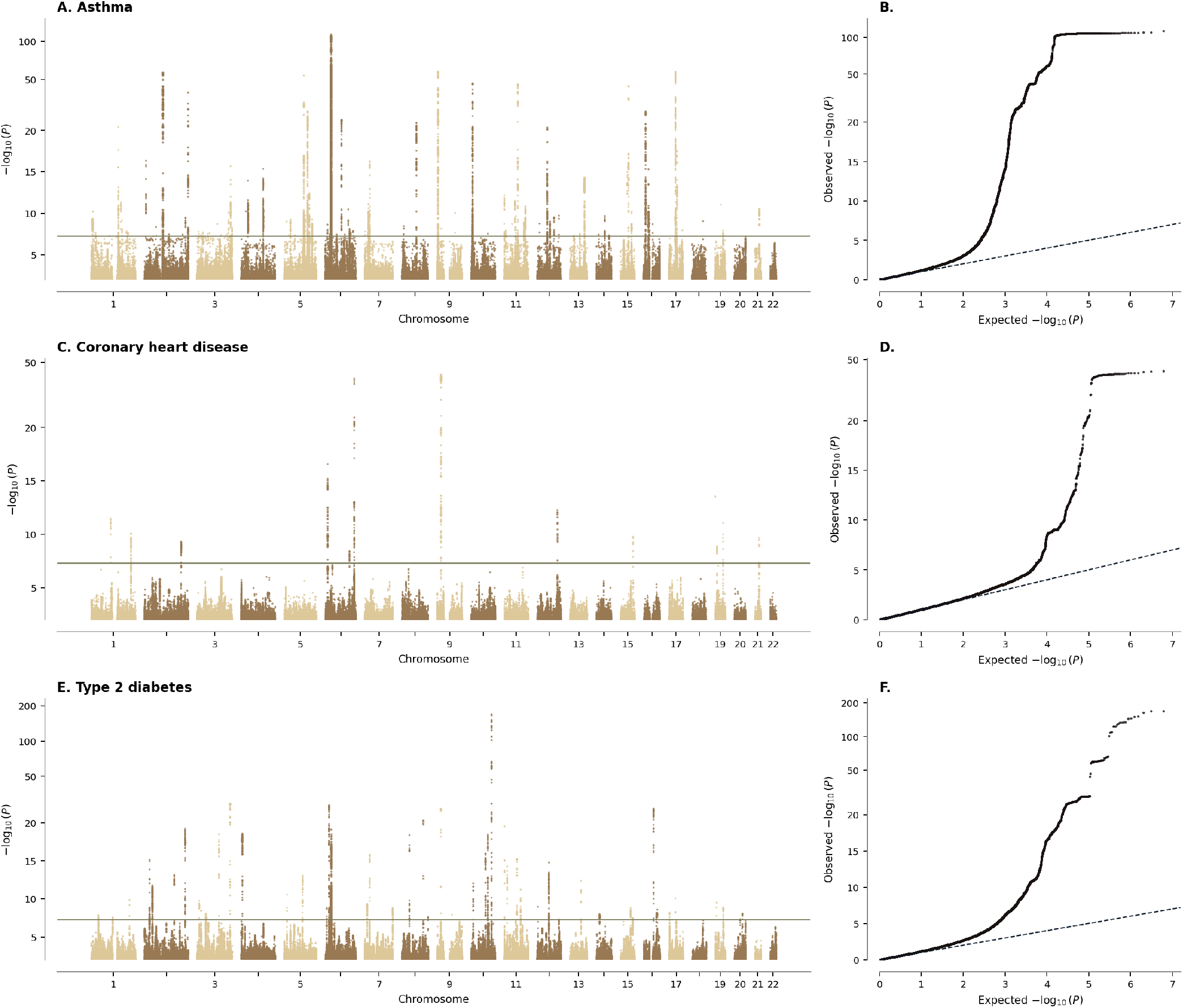
Binary-trait GWAS in the full white British cohort including related individuals (*n* = 408,624). Man-hattan (left) and quantile–quantile (right) plots are shown for asthma (**a, b**; 54,901 cases), coronary heart disease (**c, d**; 17,111 cases) and type 2 diabetes (**e, f**; 25,144 cases). Association testing residualized the 0/1 phenotype with a linear probability model and applied a logistic score test with selective saddlepoint approximation, restricted to variants with minor-allele count ≥ 20. Horizontal lines denote genome-wide significance (*P <* 5 × 10^−8^).

### 2.5 UltraSuSiE enables efficient genome-wide in-sample fine-mapping

Translating association signals into causal inference traditionally requires a distinct, computationally expensive fine-mapping workflow. Within the Cu-Gen framework, fine-mapping instead emerges as a highly efficient continuation of the primary association scan. Because UltraSuSiE directly leverages the LOCO residuals generated during the association step, the requisite sufficient statistics are already established. For each genome-wide significant locus, the algorithm utilizes the random-access capability of the .`cugen` format to rapidly extract exact in-sample covariance (**X**^⊤^ **X**) and marginal associations (**X**^⊤^ **r**). It then executes a GPU-native implementation of the Sum of Single Effects (SuSiE-RSS) model [13, 14], converging on a locus of several thousand variants in under 0.05 seconds. Crucially, by computing linkage disequilibrium directly from the analysis cohort, the resulting credible sets fundamentally eliminate the reference-panel mismatch that routinely compromises external-LD fine-mapping efforts [15].

### 2.6 Computational efficacy and scaling to multi-trait analyses

In our benchmarks, CuGen demonstrated high computational efficacy in biobank-scale analyses, with multi-phenotype runs yielding additional gains. Using standing height as a benchmark against 6.8 million imputed variants on a single high-throughput GPU with 80 GB of memory (NVIDIA H100), the array-to-imputed GWAS required 7.8 minutes in the unrelated cohort and 8.8 minutes in the full cohort (Fig. 5a). Subsequent UltraSuSiE fine-mapping of all genome-wide significant loci required 1.0 minute using eight parallel workers (Fig. 5c). Peak memory consumption remained within the 80 GB limit by streaming genotype blocks. In comparison, an identical analysis on a 16-core CPU node using REGENIE required 10.4 and 28.2 hours for the two cohorts, respectively (Fig. 5d).

**Figure 5.**
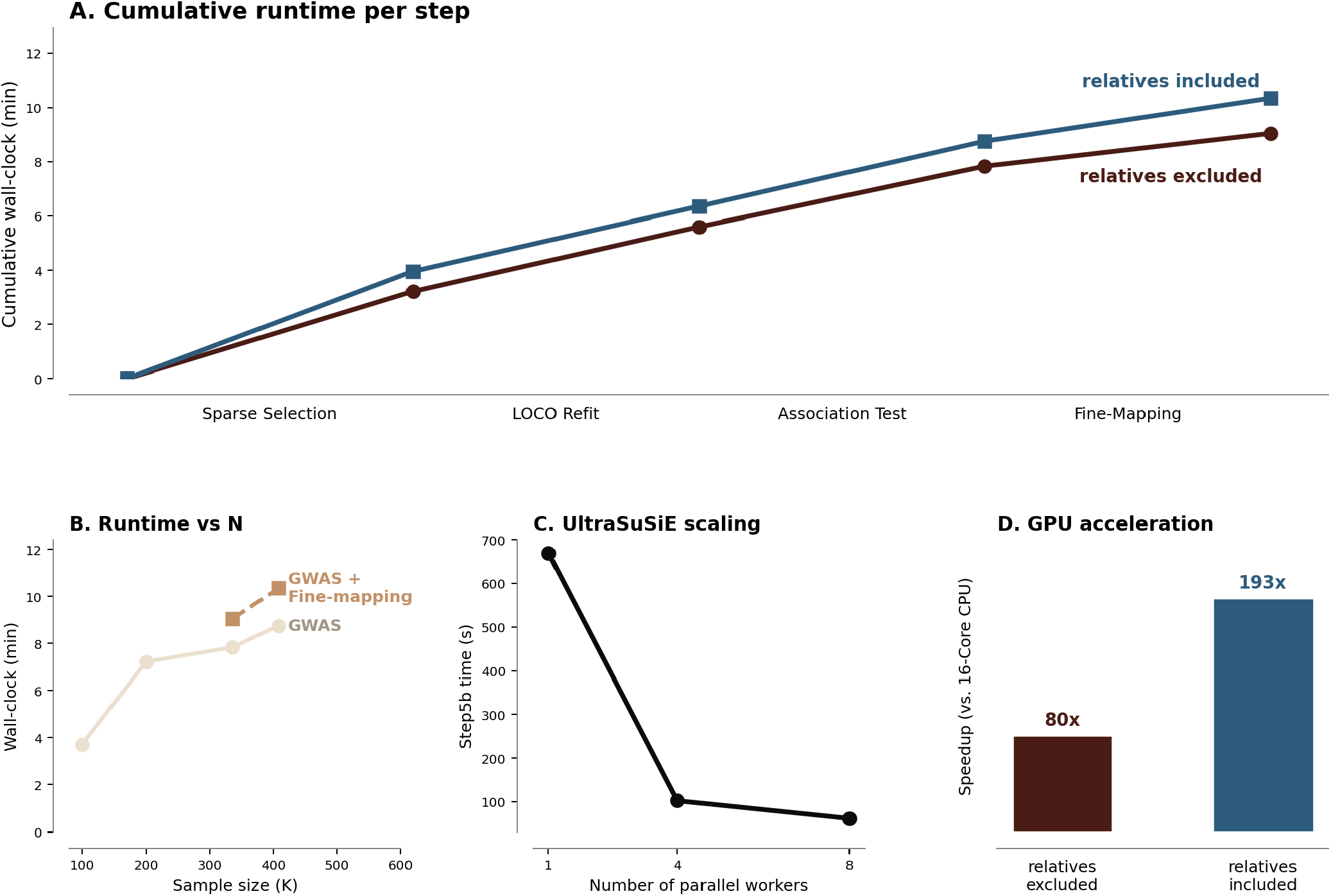
CuGen performance on standing height. **a**, Cumulative wall-clock time per pipeline step on a single NVIDIA H100 GPU, for the unrelated (*n* = 336,442) and full (*n* = 408,624) cohorts. **b**, Runtime scaling with sample size for the GWAS alone and for the GWAS with fine-mapping. **c**, UltraSuSiE fine-mapping runtime versus the number of parallel GPU workers (670 s with one worker to 62 s with eight). **d**, Speed-up over REGENIE on a 16-core CPU node: 80× for the unrelated cohort and 193× for the full cohort including relatives.

The computational efficiency increased substantially in multi-phenotype analyses. The pipeline parallelizes along two axes: the I/O-bound genome streaming stages (variant screening and association testing) fuse across multiple traits, whereas the modeling stages (joint sparse regression and fine-mapping) scale on a per-trait basis. By decoding the genotype matrix once and updating all trait accumulators simultaneously, the dominant cost of data movement is amortized across the batch. Testing an 8-trait batch on a single GPU, the full pipeline, including fine-mapping, finished in 37.7 minutes. This averaged 4.7 minutes per fully analyzed trait, demonstrating that multi-trait analyses scale sub-linearly.

CuGen’s GPU acceleration amplifies in large phenome-wide scale runs. When projected to a node of eight GPUs processing 16-trait batches, we estimated a single batch with the 6.8 million imputed variants to complete in 8.9 minutes. This achieves a processing rate of 2,579 fully analyzed phenotypes per day, averaging approximately 0.56 minutes per trait (Supplementary Fig. S7). Furthermore, the multi-trait framework scales efficiently to much higher variant counts, as only the association testing step—which most benefits from multi-phenotype processing—is affected. Increasing the number of analyzed variants to 100 million, a 16-trait batch finishes in 13.3 minutes, translating to a daily output of roughly 1,700 fully analyzed traits and averaging just 0.83 minutes per trait. Proportionally, a 1370% increase in tested variants translates to only a 50% increase in runtime (Supplementary Table S9).

### 2.7 UltraLasso inherently produces high-performance polygenic scores

The architectural design of UltraLasso inherently supports the rapid generation of highly accurate polygenic risk scores (PRS). Because the initial variant selection phases mirror the uniLasso methodology [11, 12]—utilizing leave-one-out univariate features followed by non-negative and joint LASSO selection—the resulting active set and coefficients can be deployed directly for out-of-sample prediction without LOCO residualization. When evaluated on a fixed held-out test set (*N* = 67,299), the UltraLasso-derived scores achieved predictive accuracy matching or exceeding the reference uniLasso models, approaching current state-of-the-art benchmark methods across diverse traits. Specifically, the pipeline generated highly predictive models for standing height (*R*^2^ = 0.701), body mass index (*R*^2^ = 0.115), asthma (AUC = 0.622), and coronary heart disease (AUC = 0.759) (Supplementary Fig. S6, Supplementary Table S10). Crucially, these high-performance risk scores are generated as a seamless byproduct of the GPU pipeline in a matter of minutes.

### 2.8 CuGen provides a comprehensive GPU-native genomics toolkit

To maximize the utility of the accelerated data infrastructure, CuGen extends beyond the primary GWAS pipeline to provide a comprehensive, GPU-native genomics toolkit. Leveraging the same high-throughput .`cugen` streaming reader, the framework exposes standard epidemiological operations as natively accelerated library functions. These include cohort subsetting, variant-and sample-level quality control, polygenic scoring, and the generation of Manhattan and QQ plots. By operating directly on the GPU, these routine analyses execute at interactive or near-interactive speeds. For example, processing 6.8 million imputed variants to generate per-phenotype summary visualizations takes approximately one second, while full genome-wide sample quality control completes in roughly one minute (Supplementary Table S11). By supporting this broader ecosystem of operations, the .`cugen` substrate eliminates the need for repeated I/O bottlenecks and format conversions, establishing a unified foundation for biobank-scale genetic analysis.

## 3 Discussion

The transition of biobank-scale genomic analysis from CPU-bound clusters to GPU-accelerated architectures represents a fundamental paradigm shift in genetic epidemiology [1, 2]. By demonstrating that a single GPU workstation can comfortably out-perform a multi-node CPU cluster, CuGen establishes a prelude for massive computational scaling. We condensed what traditionally requires dozens of CPU-hours into approximately ten minutes on a single NVIDIA H100 GPU. Crucially, this extreme acceleration is achieved without sacrificing statistical calibration or rigor. GPU acceleration had previously been applied to targeted genomics tasks such as expression-quantitative-trait-locus mapping [17]; CuGen extends it to the complete confounding-controlled GWAS and fine-mapping pipeline at biobank scale.

A central scientific finding of this work is the robust efficacy of sparse leave-one-chromosome-out (LOCO) prediction. Historically, controlling for population structure [18] and cryptic relatedness has relied on dense whole-genome null models [10] or mixed-model architectures [6–9]. In our previous work on uniLasso [11, 12], we demonstrated that state-of-the-art polygenic prediction can be achieved using an extremely sparse set of variants. Here, we show that this finding generalizes to confounding control: a highly compact, phenotype-informed active set—selected via hierarchical univariate-guided sparse regression— controls confounding comparably to dense ridge approaches [10] while remaining entirely GPU-memory resident. Furthermore, by selecting the candidates to maximize the explained variance of the phenotype, this sparse LOCO predictor captures substantially more post-covariate phenotypic variance. In paired simulations, this increased capture directly translated into higher statistical power at a matched false discovery rate, establishing sparse LOCO as a powerful epidemiological tool in its own right.

Furthermore, the computational speed of the CuGen framework fundamentally alters how down-stream causal inference is conducted. Currently, summary-statistics-based fine-mapping using external reference panels is widely accepted as the standard [14, 15]. However, this practice is a forced compromise driven by the prohibitive computational cost of extracting exact in-sample linkage disequilibrium (LD) at biobank scale. CuGen eliminates this limitation. By leveraging the CUDA-optimized .`cugen` genotype format, which utilizes variant-major 2-bit packing to support both high-throughput streaming and constant-time random access, UltraSuSiE executes Bayesian fine-mapping using exact in-sample LD [13, 14]. This avoids the reference-panel mismatch that frequently compromises causal variant identification [15], turning fine-mapping into an immediate, integrated extension of the primary association scan.

The practical implications of executing a biobank-scale GWAS in minutes extend well beyond raw throughput. This extreme computational efficiency drastically reduces the financial barrier to entry, democratizing large-scale multi-phenotype analyses for smaller research groups. For example, executing a scan of one hundred traits might traditionally require renting thousands of CPU cores for days, whereas the same workload can now be completed on a single H100 in a fraction of the time and cost. Additionally, this speed unlocks iterative analytical approaches that were previously prohibitive; researchers can now dynamically refine phenotype definitions, iteratively condition analyses on specific variants or environmental covariates, and perform rapid exploratory scanning in near real-time. This rapid turnaround makes the CuGen framework highly suitable for AI-assisted and agentic workflows, where subsequent analytical iterations can be orchestrated in a fully data-driven fashion [19, 20]. Finally, CuGen is engineered not merely as a standalone executable tool, but as an extensible open source GPU-native library. This modular architecture allows researchers to easily modify and build upon the existing framework, offering the flexibility required to develop novel genomic workflows.

Beyond its direct application to genetic epidemiology, the hierarchical construction of the UltraLasso predictor represents a purely statistical finding of broad relevance. We demonstrate that a LASSO [21] predictor can be successfully constructed hierarchically—screening [22] and selecting candidates locally within distinct genomic blocks before combining them into a final joint fit. The robustness of this approach is validated beyond confounding control, as our hierarchical polygenic risk scores perform on par with results derived from standard, joint-fit-only uniLasso models [11, 12]. Thus, hierarchical UltraLasso offers a novel, memory-optimized, and computationally efficient approach to statistical learning. Extending this hierarchical strategy to tackle other high-dimensional feature spaces, both in biology and beyond, remains an exciting direction for future research.

Several limitations must be addressed in future development. First, the present analyses focus exclusively on UK Biobank white British participants [23]; rigorous validation is required in admixed, founder, and non-European cohorts to ensure confounding control holds across diverse genetic architectures [24]. Second, the current pipeline utilizes hard-called imputed genotypes during model construction; future iterations must evaluate the impact of incorporating variant dosages or probabilistic encodings on power and calibration, particularly for low-frequency variants. Finally, extending viability to more widely available 32 to 40 GB GPUs through out-of-core or multi-GPU strategies remains an important objective for broader community adoption. Overall, CuGen establishes that modern genetic epidemiology software can be organized around streaming, sparsity, and the exact reuse of sufficient statistics. Rather than treating file formats and memory management as mere technical nuances, CuGen integrates them into a programmable, GPU-native library. This framework provides a unified, extensible foundation for the next generation of high-throughput, biobank-scale genomics.

## 4 Methods

### 4.1 Study population and phenotypes

We used genotype and phenotype data from UK Biobank [23]. Two white British cohorts were analyzed. The unrelated cohort contained 336,442 individuals without close relatives, and the full white British cohort contained 408,624 individuals including related individuals retained after standard quality control. Standing height (UK Biobank field INI50) was used as the primary quantitative phenotype. Binary traits were asthma (HC382), coronary heart disease (HC326), and type 2 diabetes (HC649). Quantitative phenotypes were residualized against age, sex, age^2^, sex-by-age interaction and principal components 1–10. For the binary analyses, the null model included the same covariates.

### 4.2 Genotype representation and GPU I/O

Array variants were used for sparse model construction and imputed variants were used for association testing. Array genotypes were stored in `.cugen`, a GPU-oriented binary format consisting of a header, precomputed per-variant summary statistics and 2-bit packed genotype values stored contiguously by variant. For imputed variants, best-guess hard-called genotypes were stored in a separate set of `.cugen` files; this is consistent with the approach used in REGENIE Step 2, where hard-calls are standard for common-variant association testing. The format is designed to support long sequential reads into pinned host buffers followed by asynchronous transfer to GPU memory. Read throughput for this format is reported in Supplementary Table S1.

### 4.3 Steps 1–2: univariate screening and local sparse regression

Let *x*_*j*_ denote the genotype vector for variant *j* after centering and standardization, and let *y* denote the residualized phenotype. UltraLasso first computes univariate summary statistics for streamed genotype blocks. Variants are ranked by a stability-aware strength statistic,

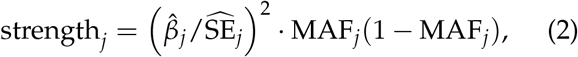

where 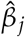 and 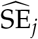 are the slope and standard error of the univariate regression of the phenotype on variant 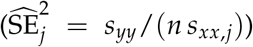. The first factor is the univariate association *χ*^2^ (squared *z*-statistic); the second, MAF_*j*_(1™MAF_*j*_), is the genotype-variance weight. Because the *χ*^2^ is invariant to rescaling the genotype, the frequency term enters exactly once, deliberately down-weighting low-frequency variants rather than double-counting frequency. Together the two factors act as a closed-form surrogate for Bayesian shrinkage at negligible cost, a frequency- and precision-weighted screen we refer to as “chainsaw shrinkage.” Both *s*_*xx*,*j*_ and MAF_*j*_ are read directly from the precomputed per-variant statistics in the .`cugen` header, so ranking the whole genome requires only the streaming univariate slope.

Within each genomic block, the top-ranked variants are passed to a local sparse regression step. This local fit is uniLasso, adopted because it attains the highest predictive accuracy per active variant [12] and therefore the sparsest active sets—the property that lets the whole model remain GPU-memory-resident. We fit a non-negative LASSO on the leave-one-out fitted values, solved by FISTA [25],

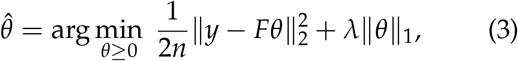

where *F* denotes the matrix of leave-one-out fitted values for the block. The leave-one-out fits in *F* carry the sign of each variant’s univariate coefficient. The non-negativity constraint on *θ* therefore rescales these univariate fits without flipping their signs—and may shrink some to exactly zero— so the composite coefficients 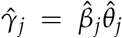 stay sign-consistent with the marginal associations. Here this construction is used only to establish the active set; the genome-wide joint LASSO and the per-chromosome OLS refit are left unconstrained.

### 4.4 Step 3: genome-wide sparse regression and LOCO prediction

Variants surviving local competition are concatenated with covariates and fit in a genome-wide LASSO, also solved by FISTA. A standard LASSO is used here rather than a further uniLasso fit: constructing uniLasso’s leave-one-out design matrix incurs a transient memory footprint far larger than the final matrix or the 2-bit-packed genotypes, so a standard LASSO maximizes the number of candidate variants that can be held in GPU memory at this stage, relying on the per-chunk uniLasso screening (Steps 1–2) to have already imposed the required sparsity. The output is a compact active set *S* that typically contains roughly 20,000–30,000 array SNPs for the quantitative analyses presented here. For each chromosome *c*, the LOCO predictor is an ordinary-least-squares refit on the active set with chromosome *c* removed,

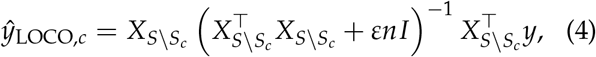

where *ε* = 10^−2^ is a small ridge. We precompute 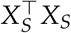 and 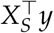 once for the full active set; each chromosome-excluded predictor is then formed by extracting the submatrix of retained columns *S* \ *S*_*c*_ and solving the corresponding normal equations directly in double precision, so the 22 refits require no further passes over the genotype matrix. This is a fresh per-chromosome solve, not a low-rank update of a single factorization. The ridge is a numerical safeguard for the occasionally near-singular Gram submatrix, not a modeling choice: on the standardized active set it imposes roughly 1% relative shrinkage, far below the L1 shrinkage already applied during selection.

### 4.5 Step 4: association testing

For a tested variant *x*_*j*_ on chromosome *c*(*j*), the LOCO residual is

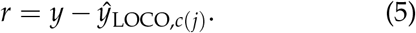

The production association statistic is a standardized regression score, formed directly from the two dot products that define the test,

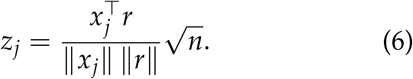

It uses the null residual variance—constant across variants, so no per-variant variance estimate is required—and matches the linear-model score statistic reported by REGENIE and related tools, making the resulting *z*-scores directly comparable. Because the test chromosome is excluded from the construction of its own LOCO predictor, the residual carries no information from that chromosome. As an alternative, a per-variant Wald statistic is available in closed form from the same dot products,

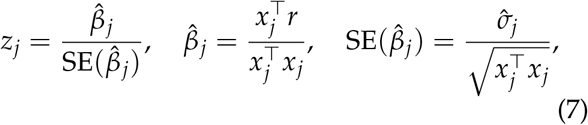

Where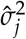 is the residual variance after fitting *x*_*j*_. On real data the two statistics are numerically almost indistinguishable (Pearson *r >* 0.9999, median |Δ*z*| = 0.000); they diverge only at the very strongest loci, where the Wald statistic is marginally larger because 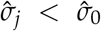 once the variant explains non-trivial variance, making the standardized score a slightly conservative equivalent (Supplementary Table S7).

### 4.6 Binary-trait implementation

For binary phenotypes, the production pipeline performs all model fitting on the linear scale and recovers logistic calibration with a single recalibration step. The 0/1 phenotype is first residualized against covariates by a linear probability model (ordinary least squares, not logistic regression), and Steps 1–3—univariate screening, local sparse regression, the genome-wide LASSO, and the per-chromosome OLS LOCO refits—are then run on this linear residual exactly as for a quantitative trait. This lets binary traits inherit the full speed of the continuous pipeline, since no logistic optimization is performed during sparse-model construction.

The linear-scale LOCO offset is then mapped back onto the probability scale by a single logistic step. For each chromosome we fit a two-parameter logistic null, logit(*µ*_*i*_) = *a* + *b η*_*i*_, where *η*_*i*_ is the LOCO offset for that chromosome, by Newton’s method; *σ*(*a* + *b η*_*i*_) recovers a calibrated null probability *µ*_*i*_. The logistic score then collapses to 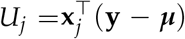, with model-based variance *V*_*j*_, computed in double precision to avoid the catastrophic cancellation that occurs when *V*_*j*_ is formed as a difference of two large near-equal sums.

Association is assessed by a score test using the normal approximation for the bulk of variants, with a Lugannani–Rice saddlepoint approximation (SPA) applied to the tail [8]. For a centered tested genotype vector *g*_*j*_, the saddlepoint calculation uses the cumulant generating function

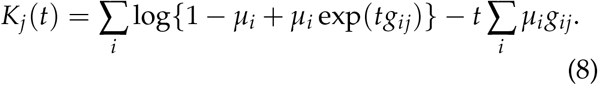

Newton iteration solves 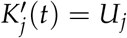, with

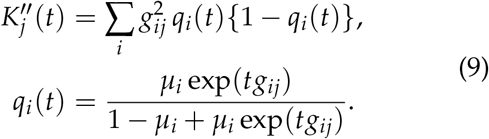

The Lugannani–Rice transform is then

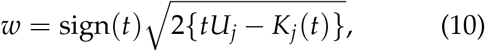

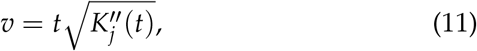

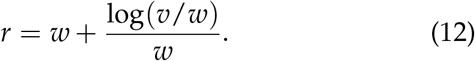

and the one-sided tail is evaluated from *r* with a sign-preserving complementary-error-function calculation; the two-sided *P*-value doubles the smaller tail. Variants for which the saddlepoint solve is invalid fall back to the normal approximation.

A variant is evaluated by SPA only if it passes the minor-allele-count floor (MAC ≥ 20) and its normal-approximation score test is at least borderline significant: *P*_norm_ *<* 0.05 and |*z*| *>* 2. All remaining variants retain the normal-approximation *P*-value. Restricting the saddlepoint evaluation to this tail set leaves the bulk *P*-values unchanged while correcting the normal approximation for low-frequency and case–control-imbalanced variants.

### 4.7 Step 5: UltraSuSiE fine-mapping

For each significant locus, UltraSuSiE computes exact in-sample sufficient statistics,

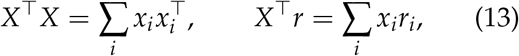

where *r* is the LOCO residual at the locus; from these it forms the in-sample LD matrix *R* (the correlation matrix of *X*) and the vector of marginal *z*-scores.

A GPU-native implementation of SuSiE-RSS [13, 14] then fits the Sum of Single Effects model to *R* and *z*. The locus effect vector is written as a sum of *L* single-effect vectors, 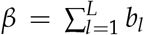 , in which each *b*_*l*_ is non-zero at a single variant only; that variant is drawn from a categorical prior *π* over the locus (uniform by default) and carries a normal effect 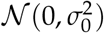. Each single effect therefore encodes “one causal variant, identity unknown,” and *L* such effects accommodate multiple independent signals at a locus. The model is fit by iterative Bayesian stepwise selection, a coordinate-ascent scheme that updates one effect at a time by a Bayesian single-variant regression on the *z*-scores residualized for the other *L* ™ 1 effects; this yields, for each effect *l*, a posterior distribution 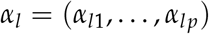 over which variant it points to, where *α*_*lj*_ is the posterior probability that effect *l* falls on variant *j* (obtained from per-variant Bayes factors). At convergence the per-variant posterior inclusion probability is

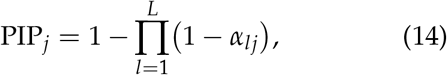

the posterior probability that variant *j* carries at least one of the *L* effects, and each effect defines a 95% credible set: the smallest set of variants whose posterior mass *α*_*l*_ sums to at least 0.95. Because *R* is computed from the same cohort that generated *z*, the fine-mapping is free from the reference-panel mismatch that affects summary-statistic fine-mapping with external LD.

### 4.8 LD Score regression

LDSC [26] was used to summarize confounding and polygenicity from GWAS summary statistics. We report the LDSC intercept, genomic inflation factor and the ratio (intercept ™ 1)/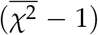 as a compact measure of residual confounding.

### 4.9 Null and power simulations

Null phenotypes were generated on real genotypes as Gaussian noise residualized against the same co-variates used in the main analyses. Polygenic simulations were generated using 1,000 causal SNPs with MAF *>* 0.01 and total heritability fixed at *h*^2^ = 0.5. Power was defined as detection of a causal locus within 500 kb at genome-wide significance, and FDR as the fraction of significant loci not within 500 kb of any true causal SNP.

To compare power directly against REGENIE, identical simulated phenotypes (10 replicates, *h*^2^ = 0.5, 1,000 causal SNPs) were analyzed by both methods in the unrelated cohort. REGENIE was run on the same array variants using its native multi-phenotype batching. Power and FDR were computed with identical 500 kb / *P <* 5 × 10^−8^ windows, and the per-replicate difference in power was assessed by a paired *t*-test and a Wilcoxon signed-rank test.

### 4.10 Sensitivity analyses

To test whether sharing a single genome-wide active set across all LOCO predictors leaks signal, we repeated the selection independently for each held-out chromosome: the candidates from chromosome *c* were dropped and the genome-wide joint LASSO re-run, yielding an active set that chromosome *c* never influenced. The resulting per-chromosome z-scores were compared to the production z-scores.

For each window (100/250/500 kb) around active SNPs, imputed variants neighbouring an active SNP were compared against a set of non-neighbours matched 1:1 on minor allele frequency and |*z*| without replacement. The UltraLasso-versus-REGENIE z-discordance (|Δ*z*|) of neighbours and matched controls was compared by a Mann–Whitney test.

### 4.11 Polygenic score evaluation

We evaluated whether the hierarchical split architecture of UltraLasso matched the predictive performance of uniLasso when used to construct polygenic risk scores. The analysis used a fixed train/validation/test split. Phenotypes were residualized within each fitting split using the same co-variates as in the association analyses. For binary traits, the phenotype was coded as a 0/1 outcome and residualized with a linear probability model.

Candidate variants were selected on the training set using the same hierarchical UltraLasso screening procedure used for GWAS model construction. Step 3 penalties were then swept on the training set, and the best penalty was selected by validation-set performance. The selected penalty was refit on the combined training and validation set. The resulting active variants and Step 3 coefficients were scored in the held-out test set.

For continuous traits, predictive performance was measured as the full-model *R*^2^ of a model including covariates and the UltraLasso score, with the covariate-only model used as the baseline. For binary traits, performance was measured as the full-model logistic AUC of a model including co-variates and the UltraLasso score. Comparisons were made against the uniLasso, Lasso, uniLasso-ES, and PRS-CS results reported previously on the same data and train/test split [12].

### 4.12 Runtime benchmarking

Runtime benchmarks were performed on a single NVIDIA H100 80 GB GPU for CuGen and on 16 CPU cores for REGENIE. Benchmarks included both sparse model construction on array variants and Step 4 association testing on imputed variants. Production runs used pinned-memory I/O and a multi-worker implementation.

Utility benchmarks were run using the same `.cugen` files and GPU-native reader infrastructure used by the main pipeline. Benchmarked operations included sample subsetting, Manhattan and QQ plotting, polygenic-score scoring, and variant- and sample-level quality control. Because CPU-native alternatives depend strongly on the specific software and hardware used, these benchmarks are reported as representative throughput demonstrations rather than as controlled cross-tool comparisons.

## Data Availability

UK Biobank data are not publicly available and were obtained through formal application to UK Biobank; this research was conducted under Application Number 24983. Access can be requested at https://www.ukbiobank.ac.uk/enable-your-research/apply-for-access. GWAS summary statistics will be deposited upon publication. The CuGen software is openly available at https://github.com/yuj1r0/cugen.

## Data availability

UK Biobank data are available through formal application to UK Biobank. GWAS summary statistics will be deposited upon publication.

## Code availability

CuGen is released as open-source software under the MIT license. The package is available at https://github.com/yuj1r0/cugen and can be installed via pip install cugen. The release includes the .`cugen` format specification and conversion utilities, the UltraLasso and UltraSuSiE implementations, and a command-line interface that supports both JSON-driven workflow orchestration and PLINK-style subcommands for common operations including quality control, polygenic risk scoring, and allele frequency computation. The current implementation is written in Python and built on CuPy [27] for GPU array computation.

CuGen is under active development as part of a broader effort to bring the standard genomics toolkit to GPU hardware. Future releases will include GPU-native implementations of population structure analysis (PCA, kinship estimation), LD computation and clumping, sample and variant subsetting, and accelerated visualization for biobank-scale datasets. The goal is to provide a self-contained GPU-native alternative to CPU-based genomics toolkits, eliminating the need to convert between formats or switch between software environments during routine analysis workflows.

## Acknowledgements

This research has been conducted using the UK Biobank Resource under Application Number 24983. M.A.R. is in part supported by the National Human Genome Research Institute (NHGRI) under award R01HG010140, and by the National Institute of Mental Health (NIMH) under award R01MH124244, both of the National Institutes of Health (NIH). T.K. was supported by the Finnish Cultural Foundation.

## Author contributions

T.K. conceived and developed the UltraLasso algorithm and the .`cugen` file format, implemented the CuGen software, performed the statistical analyses, and wrote the initial manuscript. J.R. designed and implemented the uniLasso methodology and its software for large-scale genomics. W.W. and S.L. contributed to the planning and design of the study and provided statistical and computational support. B.N., T.H., R.T. and M.A.R. provided statistical and methodological guidance, supervision, and sustained feedback that directly shaped the analyses and their interpretation. The statistical analyses reported here were designed collaboratively by all authors. M.A.R. supervised the project. All authors contributed to interpreting the results and revising the manuscript.

