## Supplementary Figures and Tables for "CuGen: A GPU-accelerated framework for large-scale genomics"

### Supplementary Information

### Supplementary Figures

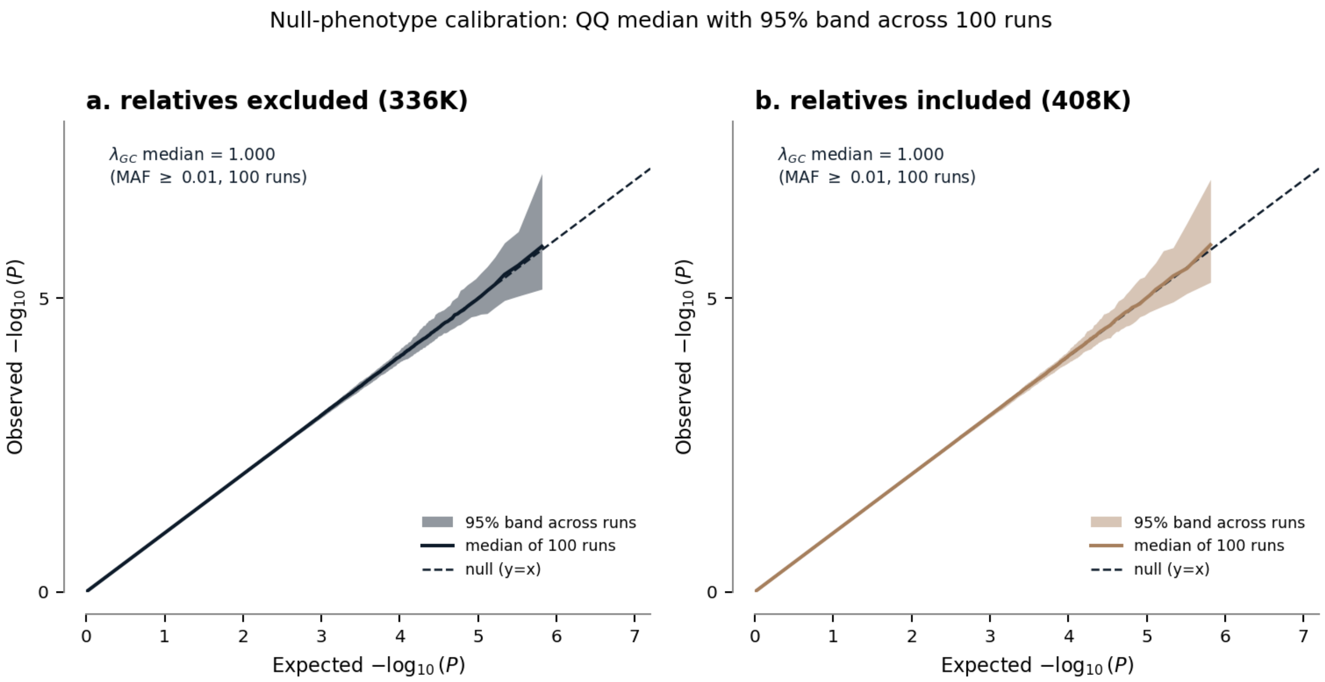


**Supplementary Fig. S1 | Null-phenotype calibration.** Quantile–quantile plots of null-simulation association statistics, showing the per-percentile median (solid line) and 95% band (shaded) across 100 null phenotypes, against the null expectation ($y = x$, dashed). **a**, Unrelated white British cohort (relatives excluded; $n = 336{,}442$). **b**, Full white British cohort (relatives included; $n = 408{,}624$). In both cohorts the median genomic inflation factor is $\lambda_{GC} = 1.000$ at common variants (minor-allele frequency $\geq 0.01$), confirming calibrated Type I error even when relatives are retained.

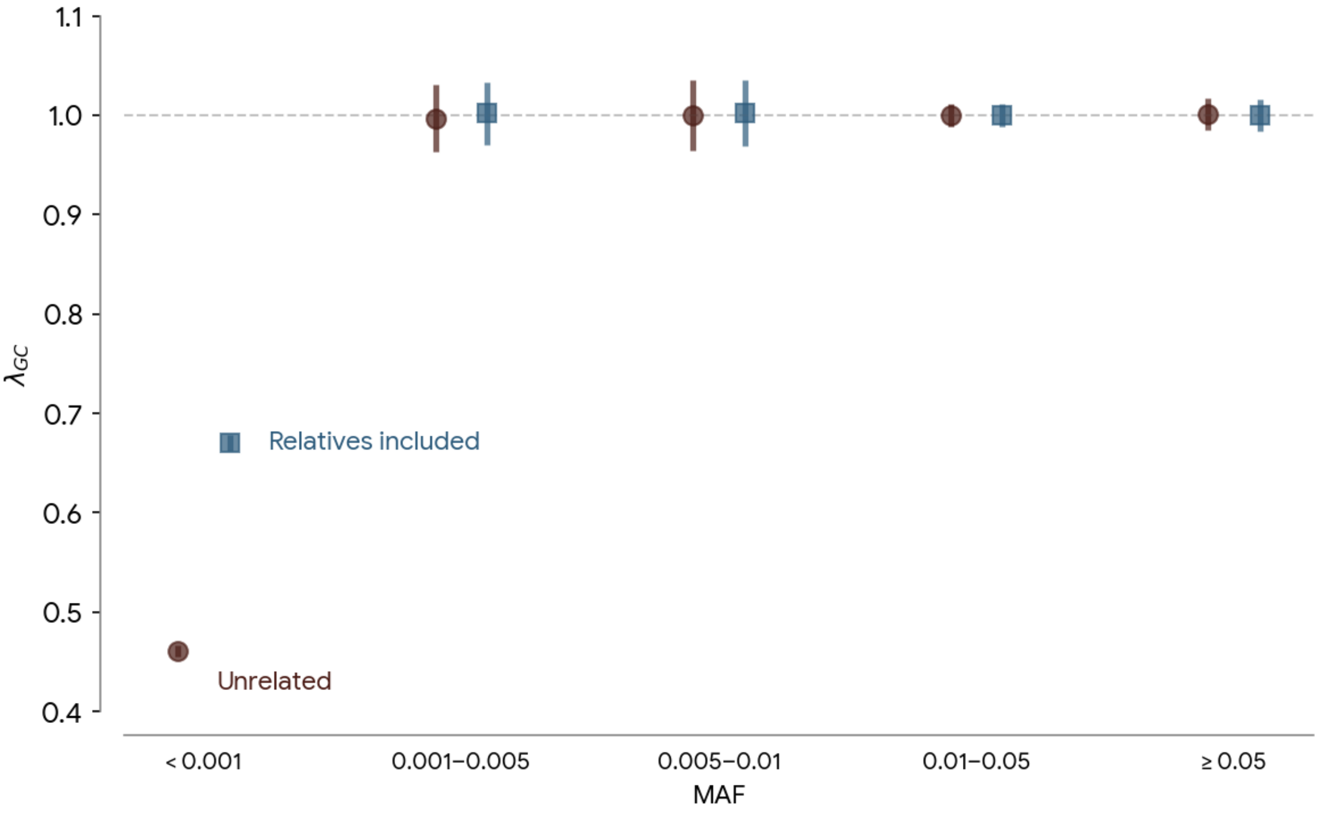


**Supplementary Fig. S2 | Genomic inflation by minor-allele-frequency stratum in null-phenotype simulations.** Genomic inflation factor $\lambda_{GC}$ as a function of the minor-allele-frequency (MAF) filter threshold for standing height. Deflation is confined to ultra-rare variants (MAF $< 0.001$), where the $\chi^2$ approximation degrades; at common variants (MAF $\geq 0.01$) inflation is at target.

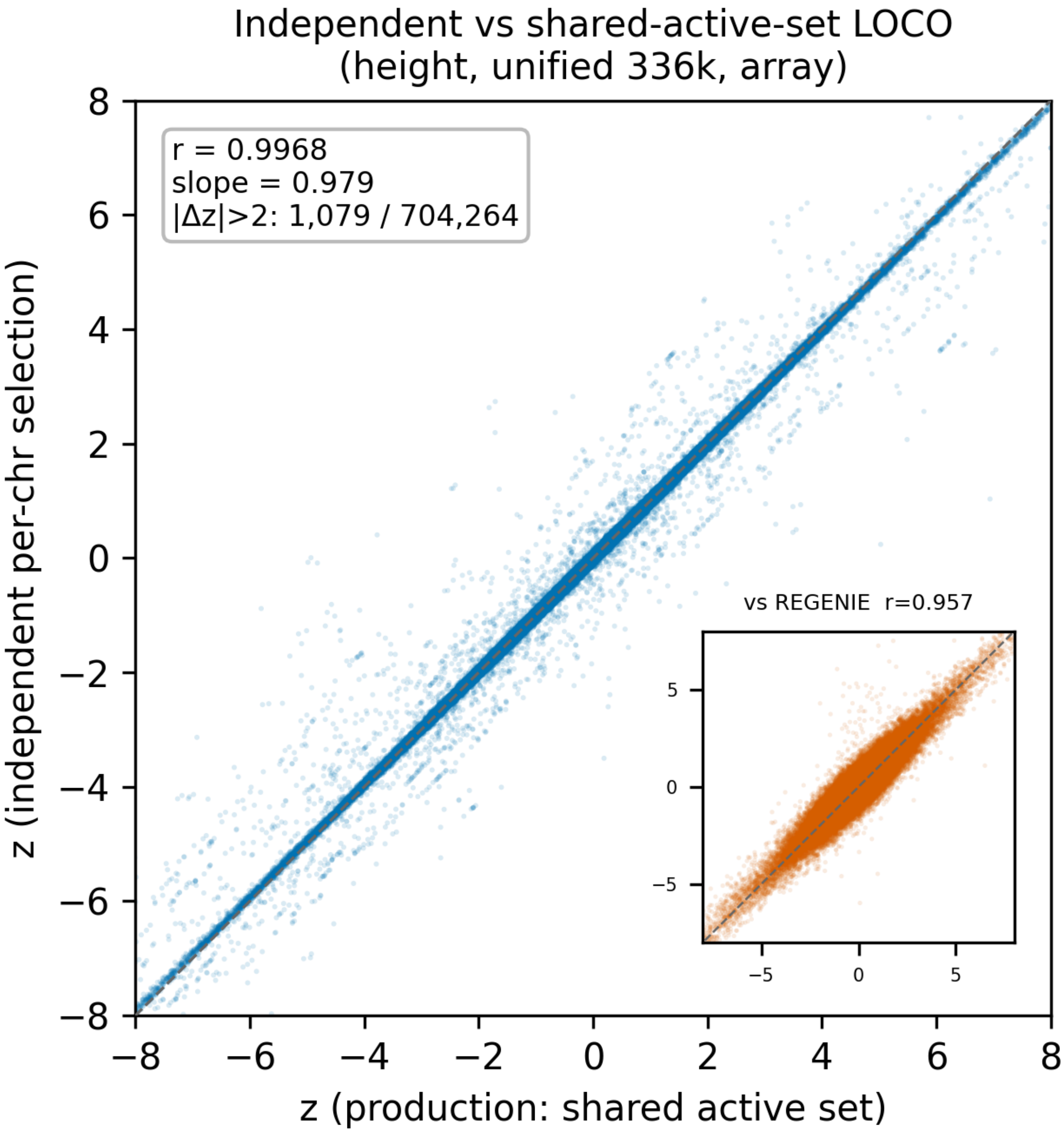


**Supplementary Fig. S3 | Independent per-chromosome selection.** Per-variant $z$-scores from fully independent per-chromosome model selection ($z_{\text{indep}}$) versus the production shared-active-set $z$-scores ($z_{\text{prod}}$) for standing height in the full cohort ($n = 408{,}624$). Pearson $r = 0.997$, slope 0.979, median $|\Delta z| = 0.03$, confirming that the single shared active set is a safe computational shortcut.

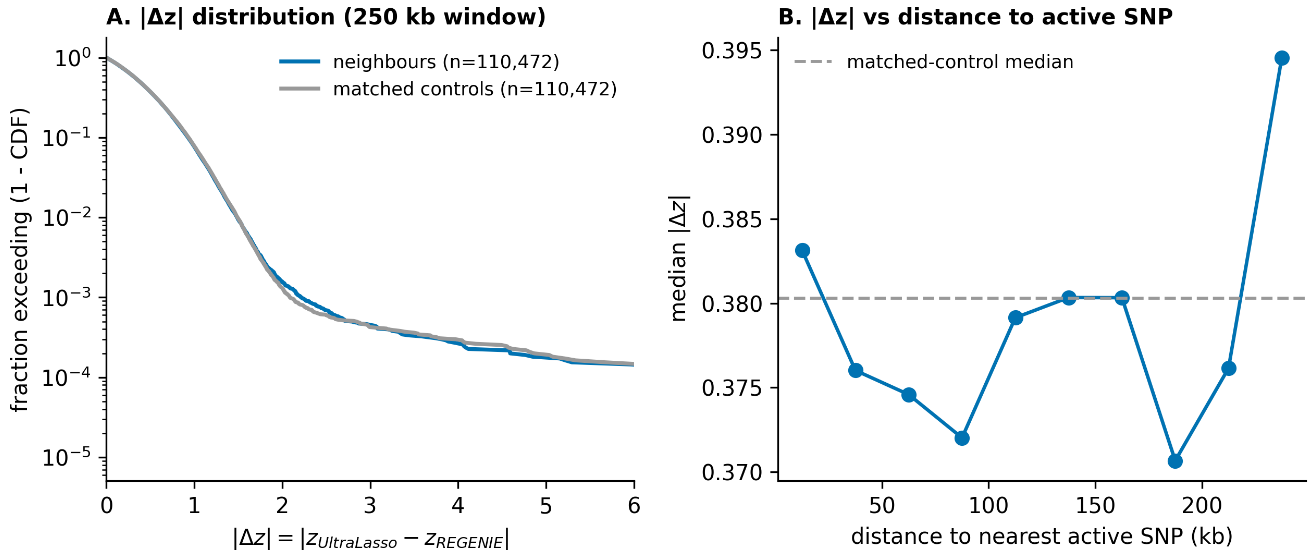


**Supplementary Fig. S4 | Neighbour $z$-score concordance.** UltraLasso-versus-REGENIE $z$-score discordance for variants neighbouring active SNPs, compared with frequency- and effect-size-matched non-neighbours, for standing height ($n = 408{,}624$). Median $|\Delta z|$ is 0.379 versus 0.380 (Mann–Whitney $P \geq 0.06$ across windows), confirming that the per-chromosome ordinary-least-squares refit removes proximal leakage.

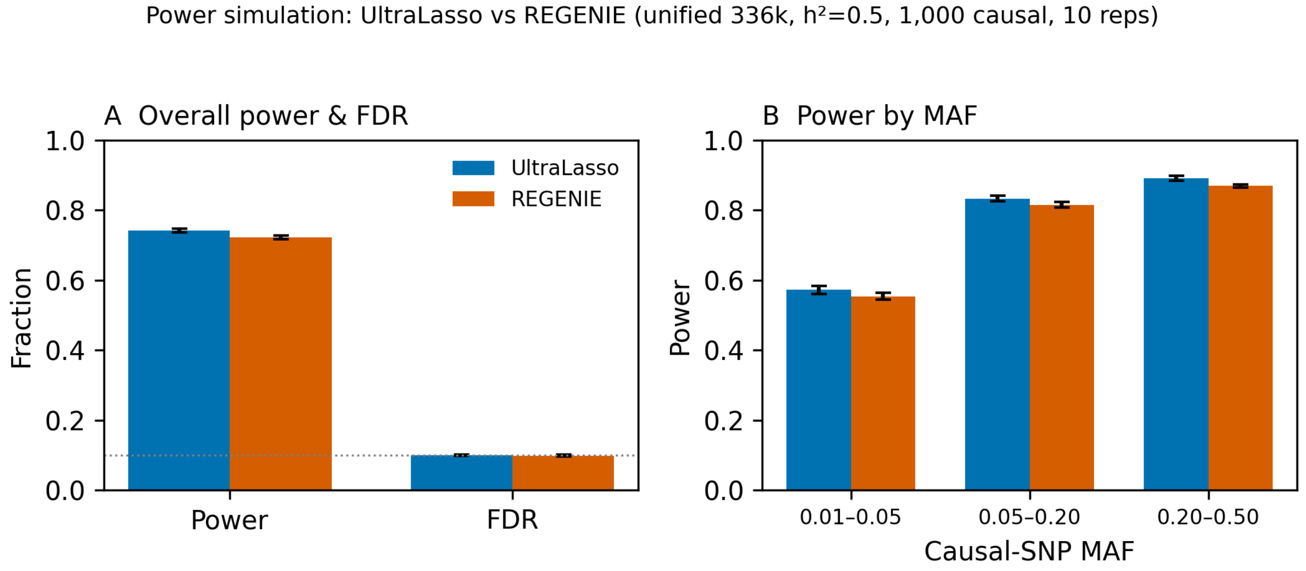


**Supplementary Fig. S5 | Statistical advantage of sparse LOCO. a**, Fraction of post-covariate residual variance captured by the LOCO predictor, UltraLasso versus REGENIE, in the unrelated ($n = 336{,}442$) and full ($n = 408{,}624$) cohorts (51.4%/49.9% versus 32.6%/34.4%, using $\sim$33$\times$ fewer predictors). **b**, Paired causal-locus power by minor-allele-frequency stratum in head-to-head polygenic simulations; UltraLasso exceeds REGENIE in every stratum (paired $t$-test $P = 3.2 \times 10^{-7}$).

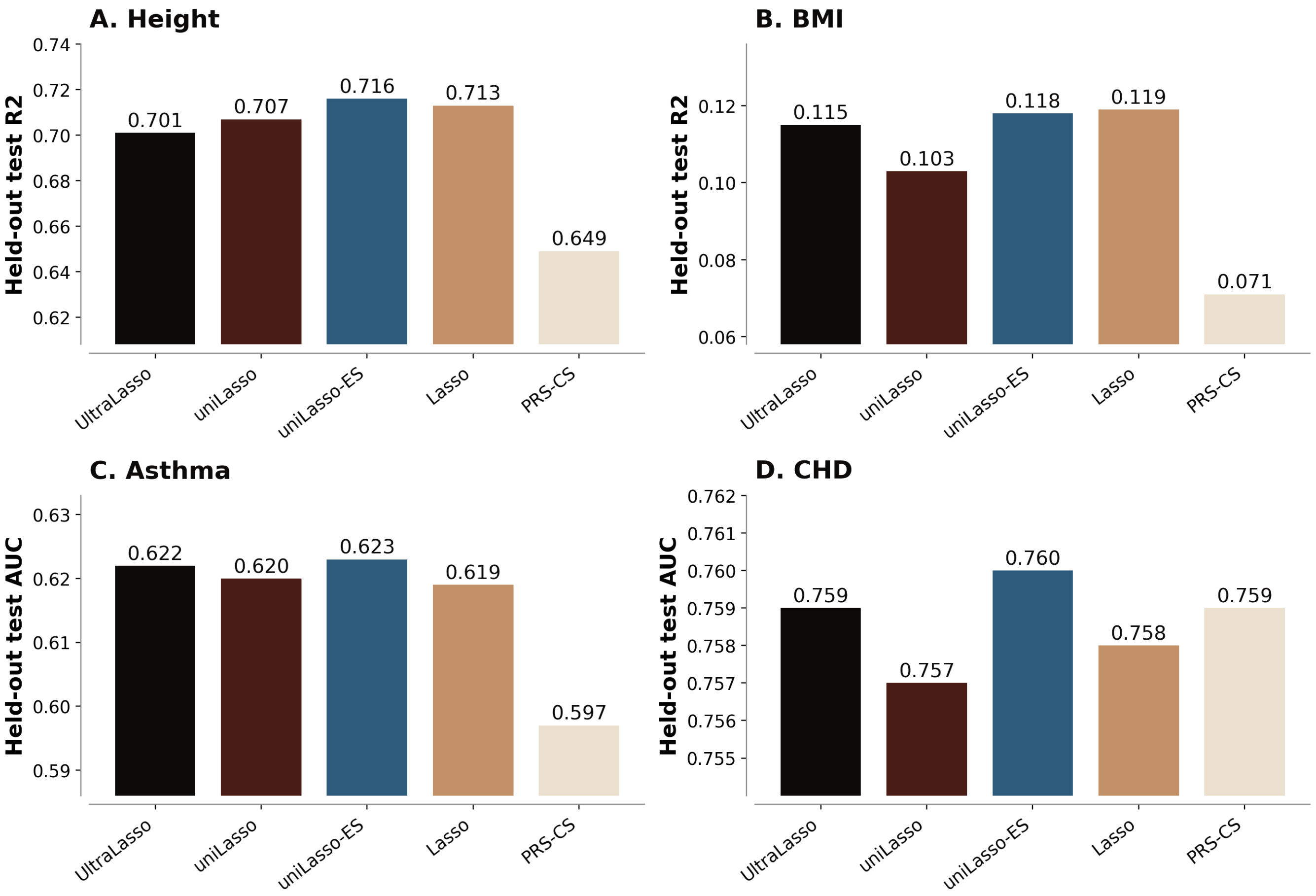


**Supplementary Fig. S6 | Polygenic-score evaluation.** Out-of-sample polygenic-score accuracy for standing height, body-mass index, asthma and coronary heart disease on a held-out test set ($n = 67{,}299$), compared with the reference uniLasso scores. Reference- and benchmark-method accuracies are taken from previously reported results on the same data and train/test split [1]. The in-sample-to-test gap (0.07 for height, 0.21 for body-mass index) quantifies the sample-specific, non-generalizable component of the in-sample fit.

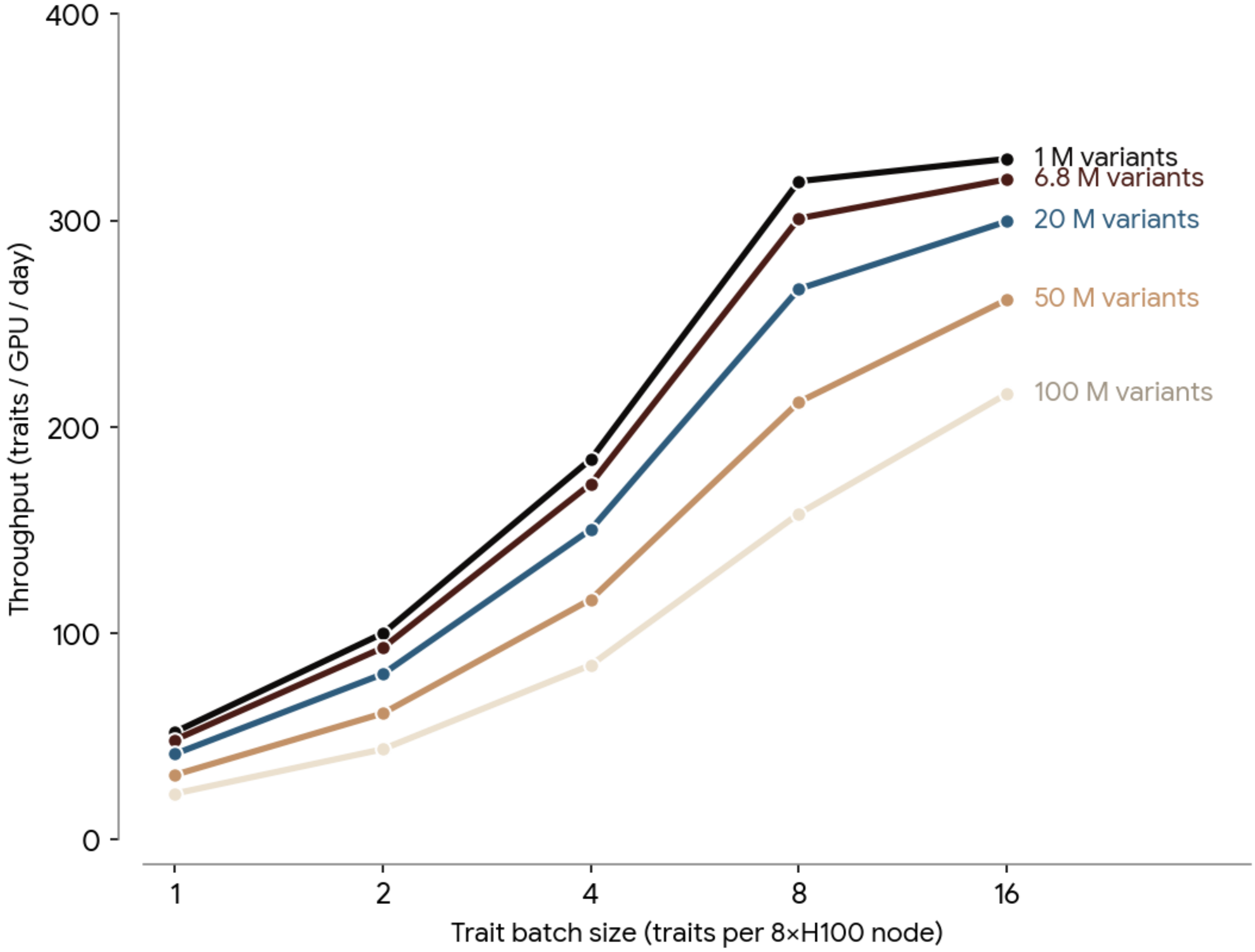


**Supplementary Fig. S7 | Multi-phenotype GPU scaling and throughput.** Full-pipeline wall-clock per trait (screening, joint LASSO and LOCO, association testing and UltraSuSiE fine-mapping) on a single 8×H100 node versus tested-variant count, for 8-trait and 16-trait batches. Because the screening and association stages fuse all phenotypes into one genome stream (the genome is read once and its I/O shared across the batch) while only the per-trait sparse-regression and fine-mapping stages scale with trait count, doubling the trait batch halves the per-trait genome-I/O slope. Filled markers, measured imputed anchor (6.82 million variants; full white British cohort, $n = 408{,}624$); shaded region, projection to whole-genome-sequencing scale. *Inset*: per-GPU throughput saturates once the trait batch reaches the GPU count ($T = G$). At 16-trait batches the node processes ∼2,600 fully-analysed phenotypes per day.

### Supplementary Table S1: Genotype read throughput

| Configuration | Reader | Median | Variants $s^{-1}$ | Speed-up |
|---|---|---|---|---|
| PLINK .pgen | Memory-mapped | 10.53 s | 1,900 | 1.0× |
| . cugen | Memory-mapped | 1.46 s | 13,673 | 7.2× |
| . cugen (1 worker) | Pinned async | 0.66 s | 30,446 | 16.0× |
| . cugen (2 workers) | Pinned async | 0.50 s | 40,069 | 21.1× |
| . cugen (4 workers) | Pinned async | **0.41 s** | **48,377** | **25.5**× |

Genotype read throughput on a single H100 SXM5 (warm cache), reading 20,000 chromosome-1 variants across all 408,624 samples into a 32.7 GB float32 matrix. Speed-up is relative to memory-mapped PLINK .pgen. Read throughput saturates near four workers, beyond which additional workers only contend for bandwidth in this compute-free benchmark.

#### Supplementary Table S2: Per-chromosome LOCO $R^2$ for standing height

| Chr | Active SNPs | LOCO $R^2$ | Chr | Active SNPs | LOCO $R^2$ |
|---|---|---|---|---|---|
| 1 | 1,953 | 0.750 | 12 | 1,298 | 0.754 |
| 2 | 1,963 | 0.750 | 13 | 553 | 0.762 |
| 3 | 1,483 | 0.754 | 14 | 677 | 0.760 |
| 4 | 1,051 | 0.756 | 15 | 826 | 0.756 |
| 5 | 1,347 | 0.754 | 16 | 884 | 0.760 |
| 6 | 1,463 | 0.750 | 17 | 972 | 0.757 |
| 7 | 1,313 | 0.756 | 18 | 488 | 0.762 |
| 8 | 1,031 | 0.758 | 19 | 760 | 0.759 |
| 9 | 1,028 | 0.757 | 20 | 638 | 0.760 |
| 10 | 1,011 | 0.758 | 21 | 274 | 0.764 |
| 11 | 961 | 0.759 | 22 | 327 | 0.764 |

In-sample leave-one-chromosome-out (LOCO) $R^2$ for standing height in the full white British cohort ($N$ = 408,624). Each LOCO predictor is an OLS refit on the active set with the listed chromosome's active variants removed; "Active SNPs" is the number of active variants residing on that chromosome. The full-genome model (all 22,301 active variants) reached an in-sample $R^2$ of 0.760, and the per-chromosome refits bracket this value (0.750–0.764), confirming that omitting any single chromosome barely perturbs the fit.

#### Supplementary Table S3: LOCO $R^2$ comparison

| Cohort | Method | $n_{\text{pred}}$ | $R^2_{\text{cov}}$ | $R^2_{\text{total}}$ | Genetics' share |
|---|---|---|---|---|---|
| Unrelated (336K) | REGENIE | 726,000 | 0.534 | 0.686 | 32.6% |
| Unrelated (336K) | UltraLasso | 22,567 | 0.534 | 0.774 | 51.4% |
| Full WB (408K) | REGENIE | 729,000 | 0.533 | 0.694 | 34.4% |
| Full WB (408K) | UltraLasso | 22,301 | 0.533 | 0.766 | 49.9% |

Genetics' share = $\Delta R^2/(1 - R^2_{\text{cov}})$, the fraction of post-covariate residual variance captured by the LOCO predictor.

#### Supplementary Table S4: Null-simulation calibration

| Cohort | $N$ | Runs | $\lambda_{\text{GC}}$ (all) | $\lambda_{\text{GC}}$ (MAF $\geq$ 0.01) | GWS FP (exp.) | Sugg. FP (exp.) |
|---|---|---|---|---|---|---|
| Unrelated | 336,442 | 100 | $0.821 \pm 0.004$ | $1.000 \pm 0.006$ | 0.02 (0.033) | 6.06 (6.53) |
| Full WB | 408,624 | 100 | $0.891 \pm 0.004$ | $1.000 \pm 0.005$ | 0.01 (0.033) | 6.43 (6.53) |

Calibration of the null-phenotype simulations: genome-wide scans of standard-normal null phenotypes (residualized on the analysis covariates) over 1,059,798 array variants, 100 null runs per cohort. Values are mean $\pm$ s.d. across runs. "GWS FP" and "Sugg. FP" are the mean number of false-positive associations per run at genome-wide ($P < 5 \times 10^{-8}$) and suggestive ($P < 1 \times 10^{-5}$) significance among common variants (MAF $\geq$ 0.01), with the value expected under calibration in parentheses. On the common set the mean genomic inflation factor is $\lambda_{\text{GC}} = 1.000$ in both cohorts and false positives are at or below nominal. The all-variant $\lambda_{\text{GC}}$ ($< 1$) reflects the expected deflation of the ultra-rare tail (Supplementary Fig. S2), not miscalibration.

#### Supplementary Table S5: Power-simulation results

| Cohort | $N$ | Reps | Power | FDR | $\lambda_{GC}$ | Power by MAF stratum $[0.01, 0.05)$ | $[0.05, 0.20)$ | $[0.20, 0.50]$ |
|---|---|---|---|---|---|---|---|---|
| Unrelated | 336,442 | 10 | $0.742 \pm 0.005$ | $0.100 \pm 0.001$ | 0.983 | 0.572 | 0.833 | 0.891 |
| Full WB | 408,624 | 10 | $0.762 \pm 0.005$ | $0.101 \pm 0.002$ | 1.081 | 0.602 | 0.851 | 0.900 |

Power and false discovery under a realistic polygenic architecture: 10 simulated phenotypes per cohort, each with 1,000 causal variants (MAF $> 0.01$) and total heritability $h^2 = 0.5$. Values are mean $\pm$ s.d. across replicates. Power is the fraction of causal loci detected within 500 kb at $P < 5 \times 10^{-8}$; FDR is the fraction of genome-wide-significant loci not within 500 kb of any true causal variant. UltraLasso recovers over 74% of causal loci at FDR $\approx$ 10% in both cohorts, with power rising monotonically across minor-allele-frequency strata. Retaining relatives (Full WB) slightly increases power without breaking FDR control; the $\lambda_{GC} > 1$ there reflects true polygenic signal ($h^2 = 0.5$), not confounding.

#### Supplementary Table S6: LDSC comparison for standing height

| Method | Cohort | $N$ | Intercept (s.e.) | $\lambda_{GC}$ | Ratio (s.e.) |
|---|---|---|---|---|---|
| UltraLasso | Unrelated | 336,442 | 1.365 (0.036) | 2.386 | 8.6% (0.9%) |
| REGENIE | Unrelated | 336,442 | 1.398 (0.036) | 2.414 | 9.4% (0.9%) |
| UltraLasso | Full WB | 408,624 | 1.374 (0.032) | 2.343 | 9.6% (0.8%) |
| REGENIE | Full WB | 408,624 | 1.503 (0.038) | 2.674 | 9.8% (0.7%) |

Array-variant summary statistics. $\lambda_{GC}$ is the genomic inflation factor (nearer 1 is better); the ratio $(\text{intercept} - 1)/(\overline{\chi^2} - 1)$ is the fraction of inflation attributable to confounding (lower is better). Reported as a comparison, not a superiority claim.

#### Supplementary Table S7: Wald versus standardized (score) association statistics

| Metric | Value |
|---|---|
| Pearson $r$ ($z$) | 0.99999997 |
| OLS slope ($z_{\text{std}}$ on $z_{\text{Wald}}$) | 0.99987 |
| Pearson $r$ ($-\log_{10} P$) | 0.9999997 |
| Median $\lvert\Delta z\rvert$ | 0.000 |
| 99.9th percentile $\lvert\Delta z\rvert$ | 0.0068 |
| Maximum $\lvert\Delta z\rvert$ | 0.1125 |
| GWS variants (both / Wald-only / std-only) | 234,738 / 29 / 0 |
| Median $\lvert z_{\text{Wald}}\rvert / \lvert z_{\text{std}}\rvert$ at GWS | 1.00006 |

Concordance between the production standardized (score) statistic and the Wald statistic on the real standing-height GWAS in the full white British cohort ($N$ = 408,624; 6,456,077 imputed variants). The two statistics are effectively identical ($r \approx 1$, slope $\approx 1$, median $\lvert\Delta z\rvert = 0.000$). The only structured difference occurs at the strongest loci, where the Wald statistic is marginally larger because the post-fit residual variance $\hat{\sigma}_j$ falls below the null variance $\hat{\sigma}_0$ once the variant explains non-trivial phenotypic variance; this yields 29 Wald-only genome-wide-significant variants versus none in the other direction. The standardized score is therefore a slightly conservative equivalent of the Wald test.

#### Supplementary Table S8: Paired power head-to-head

| Method | $n_{\text{pred}}$ | Power | FDR | MAF[.01,.05) | MAF[.05,.20) |
|---|---|---|---|---|---|
| UltraLasso | $\sim$22.6K | 0.742 | 0.100 | 0.572 | 0.833 |
| REGENIE | $\sim$726K | 0.723 | 0.099 | 0.554 | 0.816 |

Paired $\Delta$power $= +0.0192 \pm 0.0043$ (paired-$t$ $P = 3.2 \times 10^{-7}$, Wilcoxon $P = 0.002$, $n = 10$ replicates). UltraLasso detects more causal loci at every MAF stratum while holding FDR at $\sim$0.10.

#### Supplementary Table S9: Projected multi-phenotype throughput versus genome size

| | | $T = 8$ traits | | | $T = 16$ traits | | | |
|---|---|---|---|---|---|---|---|---|
| Variants | Rough regime | Step 4 | Full | Traits/day | Step 4 | Full | Traits/day | Assoc. only |
| 1 M | exome / focused set | 2.5 s | 4.5 min | 2,574 | 2.8 s | 8.7 min | 2,660 | 3,458 |
| 5 M | compact imputed GWAS | 12.5 s | 4.6 min | 2,482 | 13.9 s | 8.8 min | 2,604 | 3,364 |
| 6.8 M | current benchmark | 17.0 s | 4.7 min | 2,442 | 19.0 s | 8.9 min | 2,579 | 3,323 |
| 10 M | dense imputed GWAS | 24.9 s | 4.8 min | 2,376 | 27.9 s | 9.1 min | 2,537 | 3,254 |
| 20 M | low-frequency / WGS-ish | 49.9 s | 5.3 min | 2,188 | 55.7 s | 9.5 min | 2,414 | 3,053 |
| 50 M | dense WGS stress test | 2.1 min | 6.5 min | 1,769 | 2.3 min | 10.9 min | 2,106 | 2,577 |
| 100 M | aggressive upper stress test | 4.2 min | 8.6 min | 1,341 | 4.6 min | 13.3 min | 1,737 | 2,046 |

Projected multi-phenotype throughput on a single 8×H100 node as a function of the tested (imputed or sequenced) variant count, for 8-trait and 16-trait batches. "Step 4" is the fused association-testing time for the whole batch; "Full" is the full-pipeline batch runtime (screening, joint LASSO + LOCO, association testing and fine-mapping); "Traits/day" is the resulting full-pipeline throughput; "Assoc. only" is the throughput when fine-mapping is dropped (steps 1–4). Because only the association step scales with variant count and its genome read is shared across the batch, a 100-fold increase in variants (1–100 million) extends the full 16-trait batch runtime by only $\sim$1.5× (8.7 to 13.3 min), so throughput falls only from $\sim$2,660 to $\sim$1,740 traits/day. The 6.8-million (imputed) row is the measured benchmark; other rows are projected.

#### Supplementary Table S10: Polygenic score comparison

| Trait | Metric | UltraLasso | uniLasso | $n_{\text{active}}$ | uniLasso $n$ |
|---|---|---|---|---|---|
| Height | $R^2$ | 0.701 | 0.707 | 22,720 | 34,256 |
| BMI | $R^2$ | 0.115 | 0.103 | 21,373 | 18,833 |
| Asthma | AUC | 0.622 | 0.620 | 7,619 | 3,030 |
| CHD | AUC | 0.759 | 0.757 | 1,012 | 1,009 |

Out-of-sample test performance (held-out test set $N = 67{,}299$). UltraLasso matches or exceeds the preprint uniLasso scores on BMI, asthma and CHD, and is marginally below on height, at comparable or fewer non-zero coefficients.

#### Supplementary Table S11: Representative CuGen utility benchmarks

| Operation | Scale | Configuration | Wall-clock |
|---|---|---|---|
| Sample subset | 22 chromosomes; 81,354 of 336K samples; 21.6 GB | pinned, 4 workers | 14.1 s |
| Manhattan plot | 40 phenotypes; 6.8M imputed variants | 8 workers | 0.989 s / phenotype |
| QQ plot | 40 phenotypes; 6.8M imputed variants | 8 workers | 1.314 s / phenotype |
| PRS scoring | 732,588 weights; 336,442 samples; 22 chromosomes | single GPU | 39.8 s |
| Variant QC | chr22; 18,288 variants; 408K samples | decode + missingness + HWE | 10.14 s |
| Sample QC | 1.06M variants; 336,442 samples | single GPU | 80.4 s |

Timings were collected from verified log outputs across existing benchmark sessions and are intended to demonstrate the breadth of GPU-native operations supported by `.cugen`, not to provide a head-to-head comparison across tools. Benchmarks differ in hardware, sample size, and variant scale.

### References

[1] Joshua Richland, Tuomo Kiiskinen, William Wang, Sophia Lu, Balasubramanian Narasimhan, Trevor Hastie, Manuel Rivas, and Robert Tibshirani. Univariate-guided sparse regression for biobank-scale high-dimensional omics data. *arXiv preprint arXiv:2511.22049*, 2025.
